# Optimizing Gastric Cancer Treatment: The Role of LODDs in Lymph Node Staging

**DOI:** 10.64898/2026.02.22.26346844

**Authors:** Zhao Hao, Haiyan Niu, Yuebo Bi, Qimin Sun, Wenjun Yang

**Affiliations:** Key Laboratory of Tropical Translational Medicine of Ministry of Education, School of Basic Medicine Sciences, Hainan Medical University, Haikou, China; Department of Pathology, the First Affiliated Hospital, Hainan Medical University, Haikou, China

**Keywords:** gastric cancer, prognosis, log odds of positive lymph nodes, lymph node ratio

## Abstract

**Background:** Gastric cancer is one of the most common malignancies worldwide and is associated with poor prognosis, placing a considerable burden on public health. Overall treatment outcomes remain unsatisfactory, and accurate lymph node staging is essential for optimizing therapeutic strategies and improving survival. This study aimed to compare the prognostic value of different lymph node staging systems in patients with gastric adenocarcinoma and to provide a more refined prognostic assessment tool for clinical practice.

**Methods:** We included 4,054 patients with gastric adenocarcinoma from the SEER database (2015–2019) and 383 patients from the First Affiliated Hospital of Hainan Medical University. All patients underwent gastrectomy with D2 lymphadenectomy. Clinicopathological variables included sex, age, race, tumor size, T stage, AJCC N stage (AJCC-N), lymph node ratio (LNR), and log odds of positive lymph nodes (LODDs). Between-group comparisons were performed using the chi-square test. Optimal cut-off values were determined with X-tile software. Survival differences were evaluated by Kaplan–Meier curves. Receiver operating characteristic (ROC) curves were used to compare predictive performance. Cox regression models were applied to identify independent prognostic factors, which were then incorporated into a nomogram. Model performance was assessed using calibration curves and decision curve analysis (DCA).

**Results:** AJCC-N, LNR and LODDs were strongly and positively correlated in all three datasets (P < 0.001). ROC analysis showed that LODDs had slightly larger areas under the curve than LNR and AJCC-N for predicting 1-, 3- and 5-year survival. Multivariable Cox regression confirmed that LODDs, together with sex, age, race, T stage and tumor size, were independent risk factors for overall survival (P < 0.05). The nomogram constructed from these factors showed good agreement between predicted and observed outcomes on calibration curves, and DCA indicated meaningful clinical net benefit across a broad range of threshold probabilities.

**Conclusion:** By integrating the numbers of positive and negative lymph nodes, LODDs more sensitively reflects changes in metastatic tumor burden and showed the best prognostic performance among the evaluated systems for gastric adenocarcinoma. The proposed nomogram may serve as a useful tool for individualized prognostic assessment.

## Introduction

Gastric cancer is a major malignant tumor that threatens human health, with high incidence and mortality worldwide and a substantial impact on global public health. Epidemiological data highlight the ongoing burden of this disease. Although the incidence of gastric cancer has shown a decreasing trend in recent years, it remains the fifth most common cancer and one of the leading causes of cancer-related death worldwide. In 2022, there were approximately 968,300 newly diagnosed cases of gastric cancer and around 659,800 deaths attributable to this disease [1,2]. These figs underscore the wide prevalence and serious impact of gastric cancer on a global scale. The severity of gastric cancer is also reflected in its complex biological behavior and generally poor prognosis. Gastric cancer is highly heterogeneous, with considerable variation in clinical presentation, treatment response and survival outcomes [3]. Cancer stem cells (CSCs) are intrinsically resistant to conventional chemotherapy and radiotherapy, and they play an important role in tumor recurrence and metastasis [4]. Despite advances in diagnostic methods and treatment strategies, overall outcomes remain unsatisfactory, and the 5-year survival rate of patients with gastric cancer is still relatively low [5]. Challenges are particularly evident in patients with locally advanced or advanced disease, who often present with aggressive tumors and extensive metastasis, making treatment selection and prognostic evaluation more complex. A robust and accurate staging system is therefore crucial, as it can guide treatment decisions and support more individualized and effective therapeutic strategies.

Multiple studies have shown that different lymph node staging systems are valuable for prognostic evaluation and treatment guidance in patients with gastric cancer. For example, a hybrid lymph node staging system based on both anatomic location and the number of metastatic lymph nodes was proposed in a study of 6,025 patients who underwent gastrectomy for primary gastric cancer. That study found that, even within the same pN category, patients with higher stages in the new N classification had worse prognosis, suggesting that the anatomic extent of nodal metastasis is important and that the new system performs comparably to the current TNM classification for prognostic assessment, thus offering a possible alternative [6]. Other work comparing three lymph node staging schemes—based on the number of metastatic lymph nodes, LNR, and LODDs—in patients who received D2 resection plus adjuvant chemotherapy showed that LNR provided the best discrimination of survival among patients with locally advanced gastric cancer [7]. The number of examined lymph nodes is also critical for accurate staging and for improving survival. A cohort study of 8,696 patients with gastric cancer from the United States and China indicated that a higher number of examined lymph nodes was associated with more accurate staging and improved postoperative survival. That study suggested a minimum of 17 examined lymph nodes and an optimal number of 33 to assess the quality of nodal examination and stratify postoperative survival [8]. Another analysis of 7,620 patients who underwent curative resection for gastric cancer confirmed the importance of increasing the number of examined nodes for accurate prognostic evaluation, recommending that more than 30 lymph nodes be examined in patients with positive nodes [9]. Several newer staging systems have also shown promise. A modified pathological staging system based on LNR and the number of examined nodes has been reported to outperform the conventional AJCC staging system in prognostic assessment [10]. In addition, a hybrid system that incorporates both lymph node location and number demonstrated better prognostic performance than the 8th edition AJCC pathological N staging in a training cohort of 2,598 patients and an external validation cohort of 756 patients [11].

The log odds of positive lymph nodes (LODDs) is a relatively new method of nodal staging that has shown prognostic value in several cancers. Multiple studies have demonstrated a close association between LODDs and survival across different tumor types. For example, in a study of 440 patients with intrahepatic cholangiocarcinoma (ICC) who underwent curative resection, LODDs was an independent risk factor for survival and was proposed as the optimal lymph node-based prognostic index for ICC [12]. In patients undergoing R0 resection for pancreatic cancer, LODDs has also been identified as an independent prognostic factor, with better predictive performance than N stage and LNR among node-negative patients [13]. Compared with conventional nodal staging methods, LODDs can reduce the influence of factors related to surgery, pathology, tumor characteristics and host status on nodal assessment. For instance, when evaluating patients with adenocarcinoma of the esophagogastric junction, LODDs outperformed N stage, LNR and the number of negative lymph nodes in predicting prognosis among patients with nodal metastasis [14]. In endometrial carcinosarcoma, a prognostic nomogram based on LODDs provided more accurate and convenient prediction of overall survival than the FIGO staging system [15]. However, LODDs is not the optimal method in every cancer type. In gallbladder cancer, a prognostic model based on the number of metastatic lymph nodes showed higher accuracy than LODDs [16]. In pancreatic cancer, a staging system based on the number of metastatic lymph nodes was reported as the most suiTable prognostic indicator, with performance comparable to that of LODDs and LNR [17]. Thus, the most appropriate lymph node staging method may differ across cancer types, and the combined use of multiple indices may help optimize individualized treatment strategies and improve survival.

In this study, we used data from a Chinese clinical center in Hainan and from the SEER database, focusing on gastric adenocarcinoma. We compared lymph node staging systems from different perspectives to evaluate their impact on prognosis and to support more precise clinical decision-making.

## 1 Study Population and Methods

### 1.1 Study population

Clinical information on 24,610 patients diagnosed with gastric cancer and treated surgically between 2015 and 2019 was extracted from the SEER database. In parallel, we collected data on 418 patients who underwent surgery for gastric cancer between 2015 and 2019 at the First Affiliated Hospital of Hainan Medical University.Of 418 patients identified at our center, 35 were lost to follow-up; therefore, 383 patients were included in the external validation cohort.

Inclusion criteria were: age 18–75 years; histologically confirmed diagnosis of gastric adenocarcinoma; gastrectomy with D2 lymphadenectomy. Exclusion criteria were: palliative surgery or positive resection margins; non-adenocarcinoma histology; distant metastasis; incomplete clinical or follow-up data.

After applying these criteria, 4,054 patients with gastric adenocarcinoma from the SEER database (http://www.seer.cancer.gov) were included and randomly assigned to a training set and an internal validation set. A total of 383 patients from the First Affiliated Hospital of Hainan Medical University were included as an external validation cohort.

### 1.2 Methods

#### 1.2.1 Data collection

The following variables were collected: sex, age, race, tumor size, T stage, N stage, LNR category and LODDs category. Pathological staging followed the 8th edition AJCC TNM classification.

LNR and LODDs were defined as:

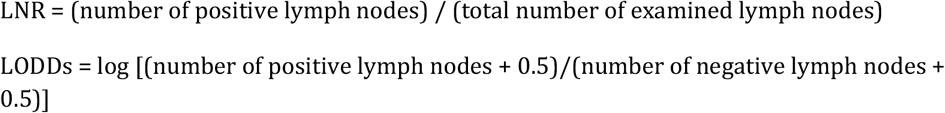

Optimal cut-off values for LNR and LODDs were determined using X-tile software (version 3.61), and these cut-offs were used to define categorical stages.

The data from the SEER database were accessed on 15/12/2024. The clinical data from the First Affiliated Hospital of Hainan Medical University were accessed for research purposes on 20/10/2024

#### 1.2.2 Follow-up

Follow-up was performed mainly through outpatient visits, telephone interviews and review of medical records or database information. Follow-up started on the date of surgery and continued until 31 December 2024. Patients were generally reviewed every 3–6 months during the first year after surgery, every 6–12 months thereafter, and once per year after 3 years of survival.

In the external validation cohort from the First Affiliated Hospital of Hainan Medical University, 35 patients were lost to follow-up, yielding a loss-to-follow-up rate of 8.37%. The last follow-up date was December 2024.

### 1.3 Statistical analysis

All statistical analyses and visualizations were performed using R software (version 4.3.2). Categorical variables were compared using the chi-square test. X-tile software was used to calculate optimal cut-off points for LNR and LODDs. Kaplan–Meier survival curves were generated to compare survival differences between groups.

Receiver operating characteristic (ROC) curve analysis and areas under the curve (AUCs) were used to compare the prognostic performance of different nodal staging systems. Variables with statistical significance in univariable analyses were entered into multivariable Cox proportional hazards models to identify independent prognostic factors.

A nomogram was constructed in R based on the results of multivariable Cox regression. Calibration curves were plotted to assess the agreement between predicted and observed outcomes. Decision curve analysis (DCA) was conducted to evaluate clinical net benefit across different threshold probabilities. A two-sided P value < 0.05 was considered statistically significant.

## 2 Results

### 2.1 Baseline characteristics

According to the inclusion and exclusion criteria, a total of 4,054 patients from the SEER cohort were included in the study. Of these, 2,838 patients were assigned to the training set, 1,216 to the internal validation set, and 383 patients from the First Affiliated Hospital of Hainan Medical University were included in the external validation set.

For all three cohorts, we summarized the baseline demographic and clinical characteristics, including age, sex, race, TNM stage (based on the 8th edition of the AJCC system), lymph node ratio (LNR), log odds of positive lymph nodes (LODDs), and tumor size (Table 1). The P-values reported in Table 1 represent statistical comparisons between the cohorts. P-values below 0.05 were considered statistically significant, indicating meaningful differences between the groups. Specifically, age, race, and TNM stage demonstrated significant variability (P < 0.001), while tumor size and N stage did not show significant differences across the datasets (P > 0.05).

**Table 1.** Comparison of baseline demographic and clinical characteristics of gastric cancer patients across the three datasets.

| Characteristics | Training set | INT Validation set | EXT Validation set | P value |
| --- | --- | --- | --- | --- |
| **n** | 2838 | 1216 | 383 |  |
| **Ages, median (IQR)** | 65 (57, 72) | 65 (56, 72) | 60 (52, 67) | **< 0.001** |
| **Sex, n (%)** |  |  |  | 0.171 |
| Male | 1768 (62.3%) | 773 (63.6%) | 257 (67.1%) |  |
| Female | 1070 (37.7%) | 443 (36.4%) | 126 (32.9%) |  |
| **Race, n (%)** |  |  |  | **< 0.001** |
| White People | 1852 (65.7%) | 800 (66.1%) | 0 (0%) |  |
| Black People | 357 (12.7%) | 131 (10.8%) | 0 (0%) |  |
| American Indian/Alaska Native | 27 (1%) | 8 (0.7%) | 0 (0%) |  |
| Asian or Pacific Islander | 582 (20.7%) | 272 (22.5%) | 383 (100%) |  |
| **TNM Stage (AJCC8) , n (%)** |  |  |  | **< 0.001** |
| 1 | 854 (30.1%) | 397 (32.6%) | 90 (23.5%) |  |
| 2 | 914 (32.2%) | 363 (29.9%) | 93 (24.3%) |  |
| 3 | 1070 (37.7%) | 456 (37.5%) | 200 (52.2%) |  |
| 4 | 0 (0%) | 0 (0%) | 0 (0%) |  |
| **T Stage, n (%)** |  |  |  | **< 0.001** |
| T1 | 724 (25.5%) | 324 (26.6%) | 76 (19.8%) |  |
| T2 | 374 (13.2%) | 157 (12.9%) | 50 (13.1%) |  |
| T3 | 1091 (38.4%) | 465 (38.2%) | 51 (13.3%) |  |
| T4 | 649 (22.9%) | 270 (22.2%) | 206 (53.8%) |  |
| **N Stage, n (%)** |  |  |  | < 0.081 |
| N0 | 1308 (46.2%) | 572 (47.0%) | 143 (37.3%) |  |
| N1 | 504 (17.8%) | 210 (17.3%) | 68 (17.8%) |  |
| N2 | 461 (16.1%) | 178 (14.6%) | 78 (20.4%) |  |
| N3a | 369 (13.0%) | 176 (14.5%) | 60 (15.7%) |  |
| N3b | 196 (6.9%) | 80 (6.6%) | 34 (8.8%) |  |
| **Tumor Size (mm) , median (IQR)** | 37 (20, 60) | 35 (20, 55) | 35 (20, 55) | 0.339 |
| **LNR, median (IQR)** | 0.041(0, 0.234) | 0.033 (0, 0.222) | 0.093 (0, 0.440) | **< 0.001** |
| **LODDs, median (IQR)** | -1.19(-1.58, -0.48) | -1.27 (-1.59, -0.52) | -0.85 (-1.46, -0.10) | **< 0.001** |

### 2.2 Correlation among the three metastatic lymph node staging systems

Spearman correlation analysis was performed in the training, internal validation and external validation sets to assess associations among AJCC-N stage, LNR categories and LODDs categories using R. In all three datasets, AJCC-N, LNR and LODDs were strongly and positively correlated with each other (P < 0.001; Fig 1).

**Fig 1.**
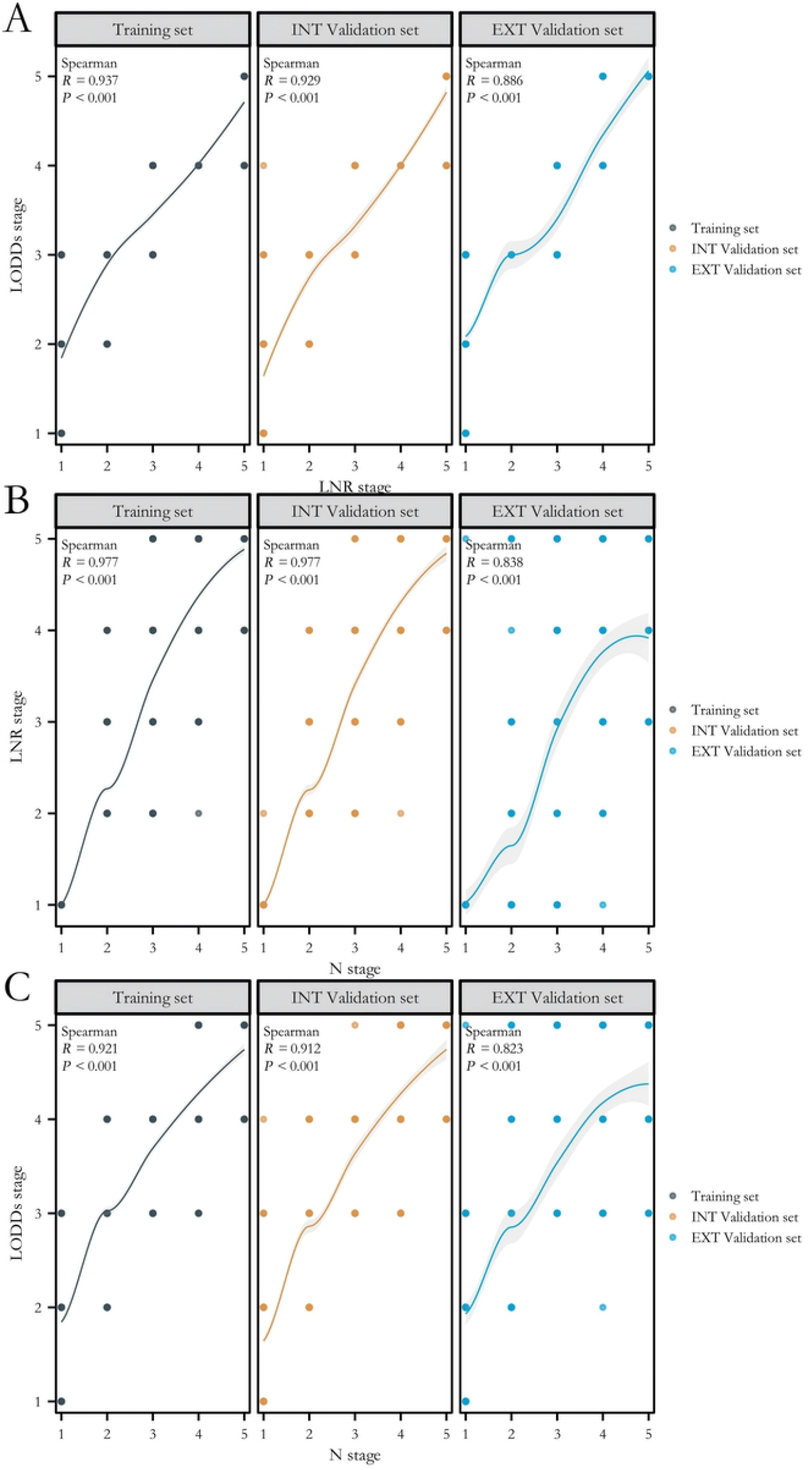
Correlation analysis among three lymph node staging methods in different datasets. (1A) LODDs vs. LNR; (1B) LNR vs. AJCC-N; (1C) LODDs vs. AJCC-N.

### 2.3 Survival curves according to the three metastatic lymph node staging systems

X-tile software was used in the training set to identify the optimal cut-off values for LNR and LODDs. The three cut-offs for LNR were 10%, 20%, 50% and 80%; the three cut-offs for LODDs were −1.8, −1.4, -0.6 and 0.2.

Based on these values, all patients were categorized accordingly. LNR was divided into five groups (LNR0, LNR1, LNR2, LNR3, LNR4) from lower to higher values. LODDs was similarly divided into five groups (LODDs0, LODDs1, LODDs2, LODDs3, LODDs4). Fig 2 shows the survival curves according to AJCC-N, LNR and LODDs in the three datasets.

**Fig 2.**
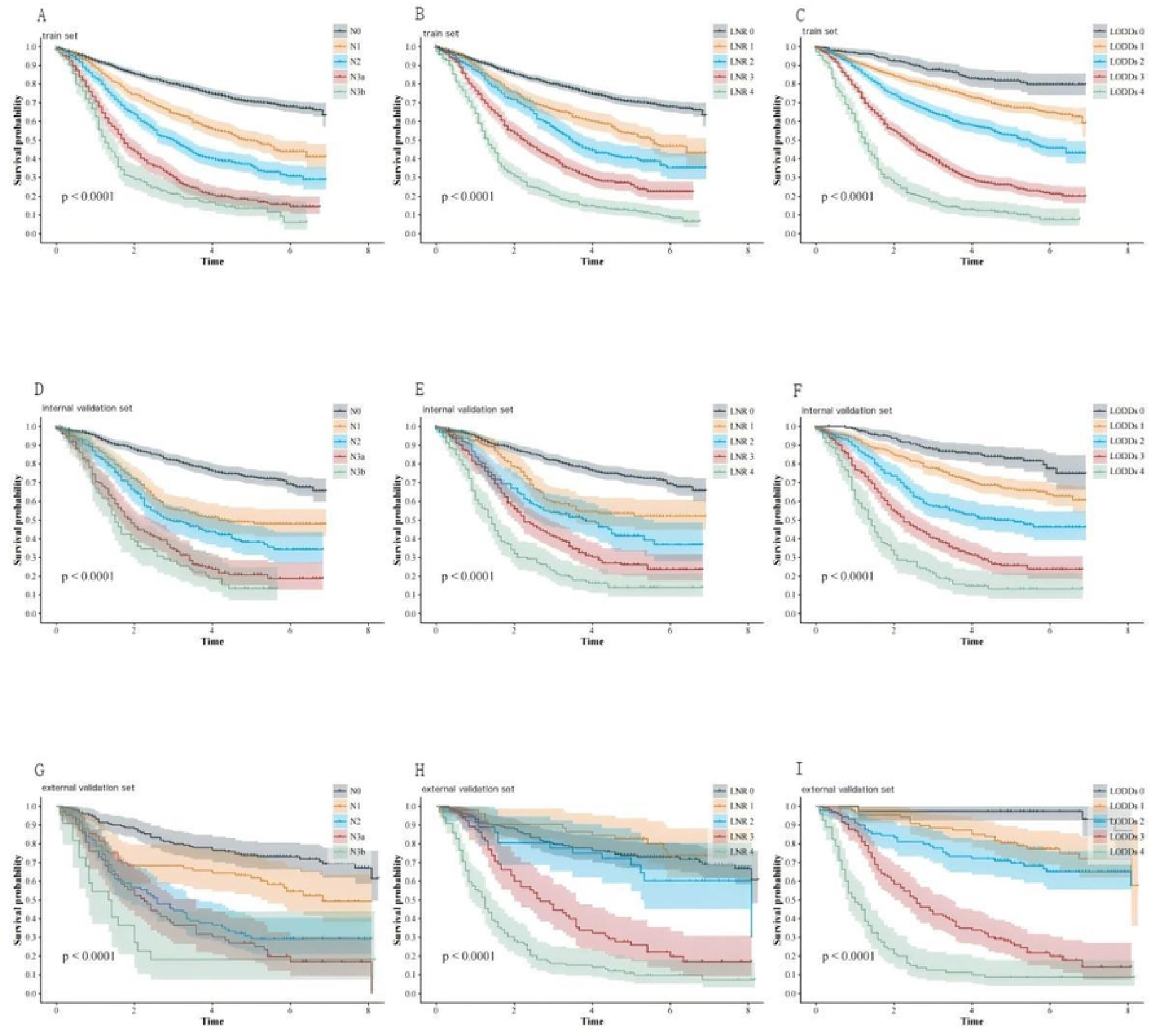
Survival curves for three lymph node staging methods in different cohorts. (2A–2C) Survival curves for the three methods in the training set. (2D–2F) Survival curves for the three methods in the internal validation set. (2G–2I) Survival curves for the three methods in the external validation set.

### 2.4 ROC curves for the three metastatic lymph node staging systems

ROC curves were plotted to compare the predictive performance of AJCC-N, LNR and LODDs for 1-, 3- and 5-year survival. In both the training and validation cohorts, the AUCs for LNR and LODDs were higher than those for AJCC-N. The AUCs for LODDs and LNR were close, with LODDs showing a slight advantage (Fig 3).

**Fig 3.**
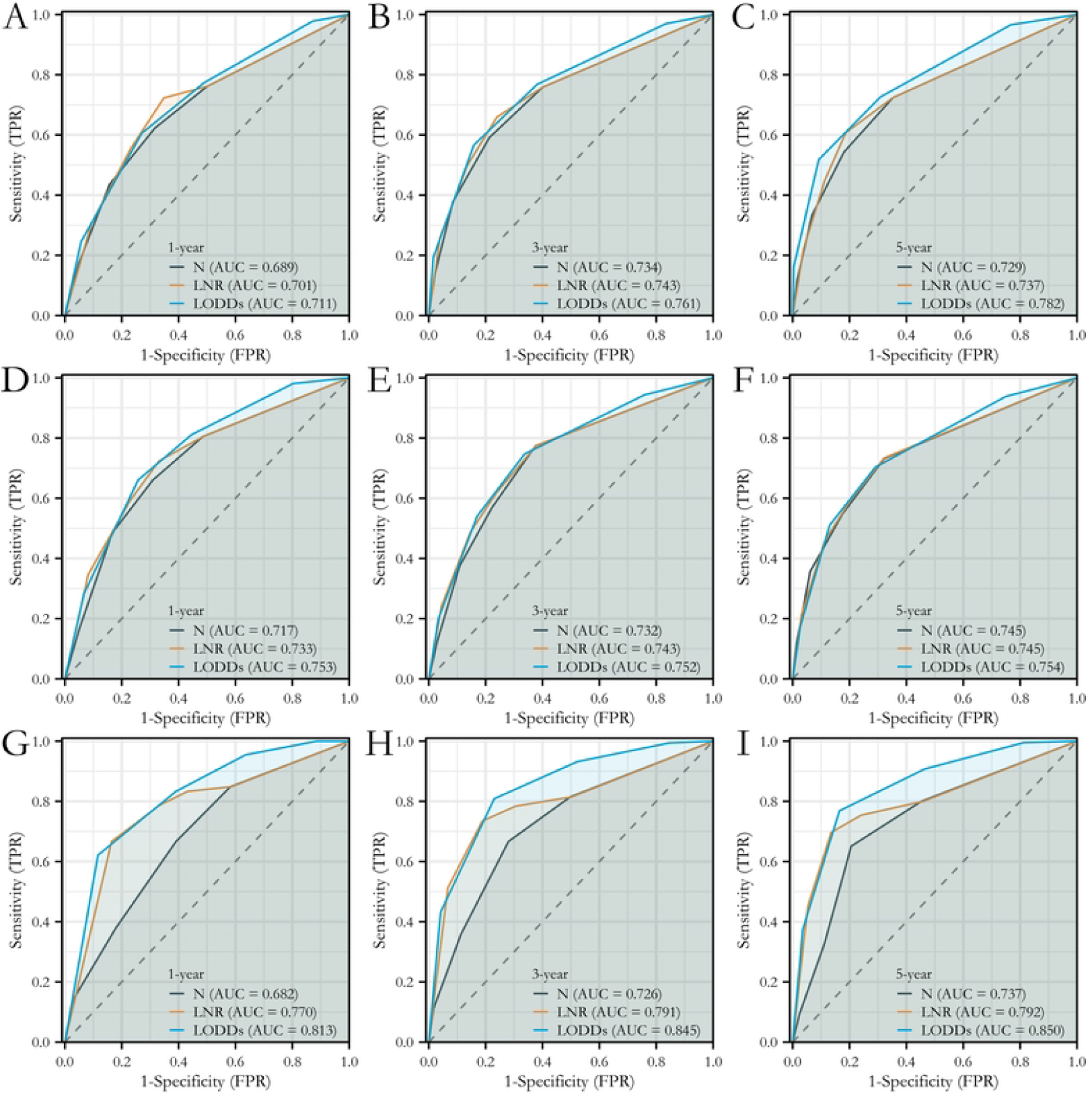
ROC curves for three lymph node staging methods across different cohorts. (3A–3C) ROC curves for 1-, 3- and 5-year survival in the training set. (3D–3F) ROC curves for 1-, 3- and 5-year survival in the internal validation set. (3G–3I) ROC curves for 1-, 3- and 5-year survival in the external validation set.

### 2.5 Univariable and multivariable analyses of overall survival in the training set

To identify independent predictors of overall survival (OS) in patients with gastric adenocarcinoma without distant metastasis, we first performed univariable analyses and then included variables with P < 0.05 in multivariable Cox regression models.

After adjustment for potential confounders, sex, age, race, T stage, LODDs and tumor size remained significantly associated with OS (P < 0.05 for all). These variables were therefore considered independent prognostic factors (Fig 4).

**Fig 4.**
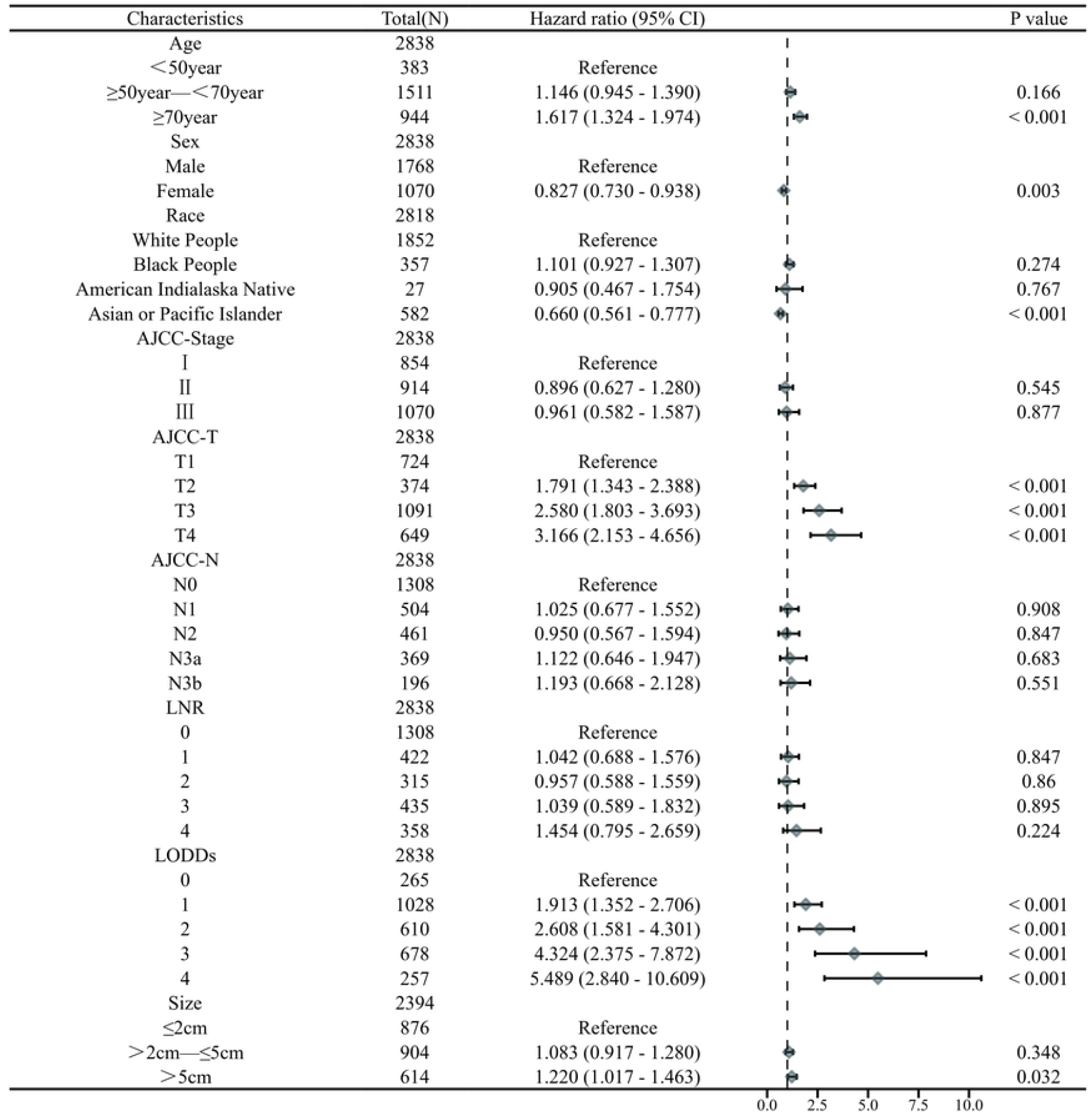
Univariable and multivariable analyses of factors associated with overall survival in the training set.

### 2.6 Construction of the nomogram and calibration curves

Based on the results of multivariable Cox regression, we used the rms package in R to calculate regression coefficients and corresponding point allocations for age, sex, race, AJCC-T stage, LODDs category and tumor size. These factors were combined to construct a nomogram that visually displays the contribution of each predictor to the estimated outcome (Fig 5A).

**Fig 5.**
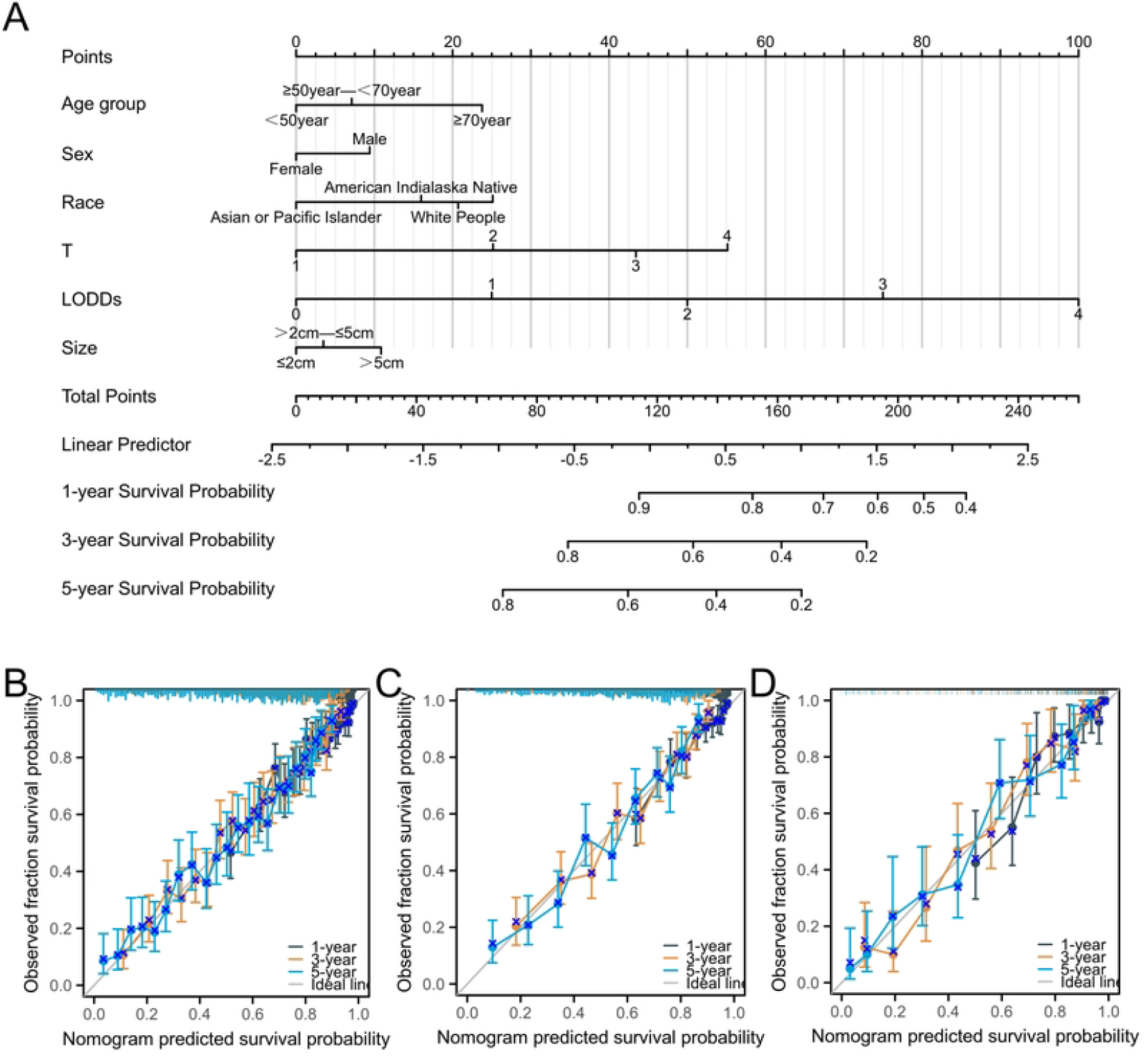
Nomogram and calibration curves based on multivariable analysis. (5A) Nomogram constructed from independent prognostic factors. (5B) Calibration curves for the training set. (5C) Calibration curves for the internal validation set. (5D) Calibration curves for the external validation set.

To evaluate the calibration of the nomogram in predicting prognosis in gastric adenocarcinoma, we generated calibration curves and used bootstrap resampling (1,000 repetitions) for internal validation to correct potential overfitting (Fig 5B–5D). The calibration curves indicated good agreement between predicted and observed survival probabilities, particularly in clinically relevant ranges. Bootstrap validation further supported the robustness of the model.

### 2.7 Decision curve analysis

To further examine the clinical utility of the nomogram, we performed decision curve analysis. DCA evaluated the net benefit associated with decisions based on the nomogram across a range of threshold probabilities (Fig 6).

**Fig 6.**
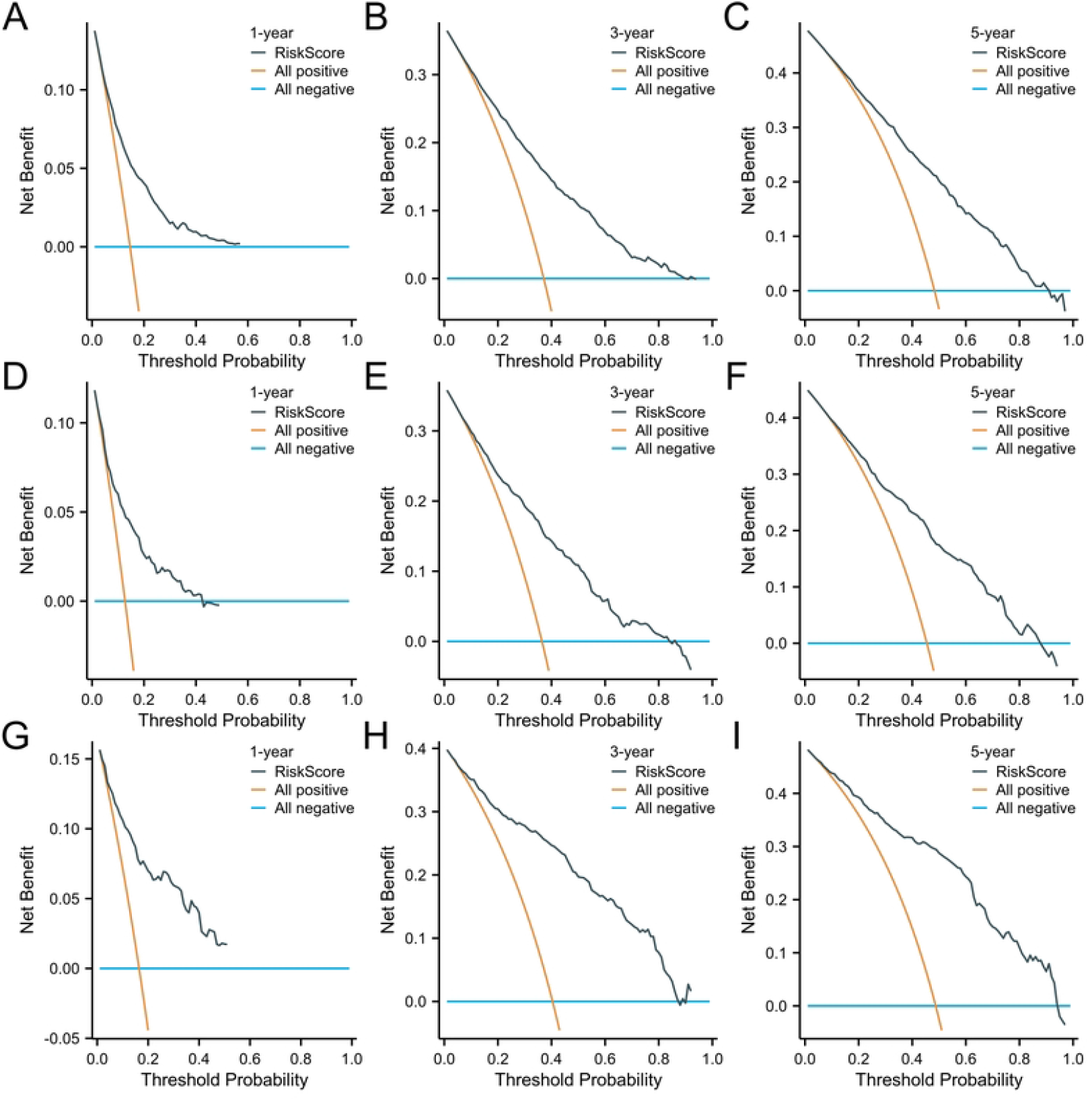
Decision curve analysis in different cohorts. (6A–C) DCA curves for the training set. (6D–F) DCA curve for the internal validation set. (6G–I) DCA curve for the external validation set.

The analyses showed that the nomogram provided positive clinical net benefit over a wide range of threshold probabilities in all three cohorts, indicating its potential usefulness in clinical decision-making.

## 3 Conclusions

By jointly considering the numbers of positive and negative lymph nodes, LODDs provides a more sensitive reflection of metastatic tumor burden than the other evaluated systems and showed superior prognostic performance in patients with gastric adenocarcinoma. The nomogram developed in this study, which incorporates LODDs together with other independent prognostic factors, may serve as an effective tool for individualized prognostic assessment.

## 4 Discussion

Gastric cancer remains one of the main causes of cancer-related death worldwide, and lymph node status is a key determinant of prognosis. Nodal metastasis is common in patients with gastric cancer and is closely associated with poorer outcomes[18]. Studies have shown that both the depth of tumor invasion and the extent of nodal involvement are important predictors of prognosis [18]. Although the AJCC TNM system, particularly the N category, is widely used for prognostic evaluation, its accuracy and discriminatory ability have been questioned [19,20]. To improve nodal staging, several alternative or complementary methods have been proposed. Among these, the metastatic lymph node ratio (LNR) has been recognized as an important prognostic factor, and several studies have reported that it provides better prognostic discrimination than the conventional pN classification [21,22].

LODDs has emerged as a promising nodal staging index and has demonstrated favorable performance in several malignancies. In gallbladder cancer, for example, LODDs-based nomograms have shown good predictive ability for 1-, 3- and 5-year survival [23]. In esophageal cancer, LODDs also demonstrated strong predictive performance [24]. Similarly, in colorectal cancer, LODDs has been validated as an independent prognostic factor [25].

In our study, we compared three approaches reflecting different perspectives: a conventional, clinically familiar system (AJCC-N), a ratio-based measure (LNR) and a mathematically transformed index (LODDs). We then integrated multiple independent predictors into a visual nomogram to provide a practical, individualized prognostic tool for clinicians. Our results suggest that LODDs offers the best discrimination among the three nodal staging systems. The nomogram that includes LODDs along with age, sex, race, T stage and tumor size demonstrated good discriminative ability and calibration in both internal and external validation cohorts. Decision curve analysis further indicated that this model may provide net clinical benefit within a wide range of reasonable threshold probabilities. The superior performance of LODDs observed in this study is consistent with previous reports. By combining the numbers of positive and negative lymph nodes using a logarithmic transformation, LODDs reduces the dependence of nodal assessment on the total number of examined nodes and may better capture subtle differences in nodal burden. Importantly, when the number of positive nodes is zero, LODDs can still differentiate patients according to the number of examined lymph nodes, whereas traditional N categories and LNR cannot distinguish among node-negative patients in this way. As a continuous variable, LODDs may therefore better reflect small changes in metastatic burden, which could underlie its improved prognostic performance.

The nomogram constructed in this study incorporates five independent predictors—age, sex, race, LODDs and tumor size—and showed better predictive accuracy than any single staging system alone. DCA suggested that the nomogram may support individualized clinical decision-making for patients with gastric adenocarcinoma without distant metastasis, help optimize allocation of medical resources and improve the cost-effectiveness of treatment strategies.

This study has several limitations. The external validation cohort was derived from a single high-volume cancer center, which may introduce selection bias. As a regional referral center, this hospital may have higher standards for surgical procedures, pathological assessment and adjuvant therapy than smaller institutions. For example, the number of examined lymph nodes may be higher than that in some primary hospitals, which could limit the generalizability of our findings, especially in settings where fewer lymph nodes are routinely examined. Further validation in multicenter, prospective cohorts with varying practice patterns would be valuable.

## Data Availability

All relevant data are within the manuscript and its Supporting Information files.

## Ethics statement

The retrospective study involving human participants was reviewed and approved by the Ethics Committee of the First Affiliated Hospital of Hainan Medical University (No. HYPLL-2025-023). In accordance with national regulations and institutional policies, informed consent was waived because anonymized archival data were used and no new interventions were introduced.

